# From Real-World Data to Virtual Intervention: A Probabilistic Neural Network for Simulating Kidney Function Preservation via Proteinuria Reduction

**DOI:** 10.64898/2026.07.12.26357786

**Authors:** Atsushi Takeda, Hideyoshi Igata, Kazue Mizuno, Yuichiro Yano, Hajime Nagasu, Mizuki Ohashi, Naoki Kashihara, Hiroyuki Kobayashi

**Affiliations:** Preferred Networks, Inc., Tokyo, Japan; Department of General Medicine, Juntendo University Faculty of Medicine, Tokyo, Japan; Artificial Intelligence Incubation Farm, Juntendo University Faculty of Medicine, Tokyo, Japan; Department of Nephrology and Hypertension, Kawasaki Medical School, Okayama, Japan; Kawasaki Geriatric Medical Center, Kawasaki Medical School, Okayama, Japan

**Keywords:** Computer Simulation, Glomerular Filtration Rate, Neural Networks, Computer, Proteinuria, Renal Insufficiency, Chronic

## Abstract

Predicting the long-term kidney function decline is critical for timely intervention but remains challenging. While the urinary protein-to-creatinine ratio (uPCR) is a potential surrogate endpoint, its short-term reduction’s link to long-term nephroprotection requires investigation. This study aimed to develop a probabilistic neural network model to capture both the estimated glomerular filtration rate (eGFR) slope and its uncertainty based on baseline clinical characteristics. Using a retrospective dataset, we designed a neural network to output a predictive distribution (mean μ and standard deviation σ) for the eGFR slope. SHAP (SHapley Additive exPlanations) was used for model interpretation, and a simulation study quantified the impact of uPCR reduction. In the validation set, the model achieved a Pearson’s correlation coefficient of 0.56 and an RMSE of 2.81 ml/min/1.73m²/year between predicted and actual slopes. SHAP analysis identified uPCR as the most potent predictor, with higher baseline levels associated with a more rapid eGFR decline. Furthermore, a simulated 62% uPCR reduction demonstrated a significant improvement in the predicted eGFR slope, an effect most pronounced in patients with high baseline uPCR. This proof-of-concept study reinforces the critical role of uPCR in predicting eGFR slope and suggests its reduction may contribute to long-term kidney function preservation, warranting validation in larger, diverse real-world datasets.

## Introduction

The progression of chronic kidney disease (CKD) is a growing global health concern, particularly due to its asymptomatic early stages and the limited effectiveness of treatment options once significant renal function has been lost.^1–4^ Predicting long-term kidney function decline is critical for identifying patients at high risk and initiating timely intervention.^5–7^ However, estimating the future slope of the estimated glomerular filtration rate (eGFR) remains a formidable challenge, especially when relying solely on baseline clinical parameters.^8^

Among potential biomarkers, the urinary protein-to-creatinine ratio (uPCR) has garnered attention as a surrogate endpoint in nephrology. Several studies have demonstrated the prognostic significance of proteinuria reduction in slowing CKD progression.^9–13^ However, it remains uncertain whether a short-term reduction in uPCR translates into long-term eGFR outcomes across a diverse population of CKD patients. A key question is whether short-term reductions in proteinuria, such as those induced by pharmacologic interventions, reflect genuine nephroprotection or merely transient hemodynamic effects.^14–15^

In this study, we aimed to address the challenge of forecasting long-term kidney function decline by developing a probabilistic neural network model that captures both the expected trajectory and the uncertainty of the eGFR slope. Rather than relying on single-point predictions, we trained the model to learn a full probability distribution of the eGFR slope based on baseline clinical characteristics.

To interpret the model and identify the most influential predictors, we employed the SHapley Additive exPlanations (SHAP) framework.^16^ Based on the SHAP analysis, we conducted a simulation study to evaluate the effect of reducing uPCR, which was identified as a particularly important predictor, on the predicted rate of eGFR decline. By modeling the relationship between the degree of proteinuria reduction and kidney function preservation, we sought to provide a data-driven basis for evaluating uPCR as a modifiable target and surrogate endpoint. This work should be viewed as a proof-of-concept study that demonstrates the feasibility of this modeling approach and provides a foundation for future research in real-world datasets.

## Materials and Methods

### Ethical Approval and Consent to Participate

This study was a secondary analysis of publicly available data from the “Prognosis of chronic kidney disease with normal-range proteinuria: The CKD-ROUTE study”.^17^ The original study (UMIN Clinical Trials Registry: UMIN000004461) was approved by the ethical committees of Tokyo Medical and Dental University, School of Medicine (No. 883) and all institutions participating in the study and was conducted in accordance with the ethical principles of the Declaration of Helsinki.

The protocol for this present study was reviewed and approved by the Institutional Review Board of the authors’ institution. As the study utilized publicly available and de-identified data, according to the “Ethical Guidelines for Medical and Biological Research Involving Human Subjects” published by Japanese government and the ethical review regulations of the authors’ institution, the need for additional participant consent was waived.

### Dataset and Study Population

We performed a retrospective analysis using a publicly available dataset from Iimori et al. (2018).^17^ At baseline (month 0), several clinical characteristics were recorded, of which we used 16 variables for our analysis (Table1).^5,18^ In the original study, eGFR was measured every 6 months for a maximum follow-up period of 36 months.

### Outcome Definition and Cohort Selection

The primary outcome for our analysis was the eGFR slope, which represents the rate of kidney function decline. From the total of 1,138 samples in the dataset, we first selected patients who had five or more eGFR measurements (n = 655).^19^ Then, samples with missing values in any of the 16 features (n = 115) were excluded from analysis. For the remaining individuals, the eGFR slope was calculated using an ordinary least-squares linear regression model. From this cohort, we then excluded patients (n = 4) whose absolute eGFR slope was ≥15 ml/min/1.73m²/year, as these values were considered clinical outliers. In total, 536 samples were included in the following analyses.

### Data Preprocessing

The raw dataset underwent a series of preprocessing steps to prepare it for the model.

1. Categorical Data Encoding: Non-numeric categorical features were transformed into a numerical format using one-hot encoding. To avoid multicollinearity, the first category of each feature was dropped.
2. Feature Engineering: To capture potential non-linear relationships, a new feature was engineered from the existing uPCR feature. Specifically, the natural logarithm of the uPCR value, log(uPCR+ɛ), was computed and appended to the feature set. A small constant, ɛ=0.005, was added to ensure the logarithm’s argument was positive.
3. Dataset Splitting: The preprocessed dataset (536 samples) was randomly partitioned into a training set (80%; 428 samples) and a validation set (20%; 108 samples). The training set was used for model learning, while the validation set was reserved for model evaluation.

### Model Architecture

We designed a neural network that outputs the parameters of a Gaussian (normal) distribution for each input instance.

1. Input Standardization: All input features were standardized by subtracting the mean and dividing by the standard deviation. These scaling parameters (μ_scaler_ and σ_scaler_) were computed exclusively from the training data and stored within the model. The standardization is defined as:

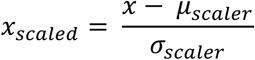
2. Feature Extractor Network: The core of our model is a feed-forward neural network consisting of two hidden layers, each with 64 neurons and a Rectified Linear Unit (ReLU) activation function. This network processes the standardized input vector, *x_scaled_*, to generate a latent feature representation, *f*.
3. Probabilistic Output Heads: The latent feature vector, *f*, is passed to two independent linear layers, or “heads,” to predict the two parameters of a Gaussian distribution:

- Mean (μ) Head: A linear layer that outputs the predicted mean of the distribution.

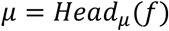
- Standard Deviation (σ) Head: A linear layer whose output is transformed to ensure the standard deviation is always positive. This is achieved by applying the softplus function, and a small constant (*ε*_σ_ = 1.0 × 10^−6^) is added for numerical stability during training.

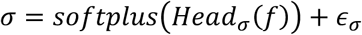

The final output of the model for a given input is the parameter pair (μ,σ), which defines a complete predictive probability distribution.

### Model Training and Evaluation

The model’s parameters were optimized by maximizing the likelihood of the training data given the predicted distributions.

1. Loss Function: We used the Negative Log-Likelihood (NLL) of the Gaussian distribution as the loss function. For a true target value *y_true_* and predicted parameters (μ,σ), the loss L is calculated as:

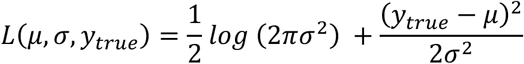

Minimizing this loss function is equivalent to maximizing the log-likelihood of the observed data.
2. Optimization: The model was trained using the Adam optimizer with a learning rate of 0.0001.
3. Early Stopping: To prevent overfitting, an **e**arly stopping mechanism was implemented. We monitored the Root Mean Squared Error (RMSE) on the validation set at the end of each epoch. If the validation RMSE did not improve for 10 consecutive epochs, the training process was terminated. The model weights from the epoch with the lowest validation RMSE were selected as the final model.

### Model Interpretability and Feature Analysis

To understand the behavior of the trained model and to identify the key drivers of its predictions, we employed the SHAP (SHapley Additive exPlanations) framework.^16^ SHAP provides a unified and theoretically sound method to explain the output of any machine learning model by assigning each feature an importance value for a particular prediction.

1. Explainer Configuration: Since our model outputs two parameters (the mean μ and the standard deviation σ), and our primary interest lies in explaining the model’s central prediction, we created a wrapper model. This wrapper isolates the model’s forward pass to return only the predicted mean, μ. We utilized the GradientExplainer from the SHAP library, which is specifically designed for deep learning models. The explainer was initialized using the entire training dataset as the background distribution to establish a baseline for calculating the Shapley values.
2. SHAP Value Calculation: With the configured explainer, we computed the SHAP values for each feature for every instance in the validation set. These values quantify the positive or negative contribution of each feature to the model’s prediction of μ for that specific instance.
3. Visualization for Interpretation: The computed SHAP values were used to generate several visualizations to interpret the model’s behavior at both global and local levels:

- Global Feature Importance: We generated SHAP summary plots to understand the overall impact of each feature. The dot plot visualizes the distribution of SHAP values for each feature, indicating not only its importance but also the direction of its correlation with the prediction. A bar plot was also used to rank features by their mean absolute SHAP value, providing a clear overview of the most influential features.
- Feature Dependence: SHAP dependence plots were created for key features (e.g., “uPCR”) to visualize how the value of a single feature affects the model’s output across the dataset. These plots can also reveal potential interaction effects with other features.
- Individual Prediction Explanation: To explain specific predictions, we generated SHAP waterfall plots. These plots deconstruct a single prediction, illustrating how each feature’s positive or negative contribution pushes the model’s output from the base value (the average prediction over the training set) to the final predicted value for that instance. This allows for a detailed, transparent view of the model’s reasoning for individual cases.

### Simulating the Clinical Impact of uPCR Reduction

To quantify the potential clinical benefit of reducing the uPCR on the eGFR slope, we conducted a simulation study using the trained model. This analysis aimed to model the effect of a therapeutic intervention on a representative patient cohort.

1. Creation of a Synthetic Patient Cohort: We first constructed a synthetic dataset representing a cohort of “average” patients (Table 2). A baseline patient profile was established by taking the mean value for all continuous features and the modal (most frequent) value for all categorical features from the original training dataset. Based on this average profile, we generated 100 synthetic patient instances where all features were held constant, except for the uPCR value, which was varied logarithmically from 0.01 to 10.0. This process created a controlled cohort of patients differing only in their initial uPCR level.
2. Counterfactual Simulation: We simulated a clinical intervention corresponding to a 62% reduction in UPCR.^20^ For each patient in the synthetic cohort, we generated a “post-intervention” counterfactual profile by applying this reduction to their uPCR value. The model was then used to predict the eGFR slope distribution for both the original 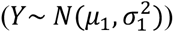 and the post-intervention 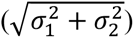 profiles.
3. Quantifying the Improvement: The effect of the intervention was quantified by analyzing the distribution of the *change* in the eGFR slope, defined as *ΔY* = *Y*^′^ − *Y*. As the difference between two independent normal distributions, the distribution of this change is also normal:

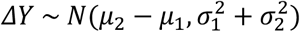

Using this derived distribution, we calculated the expected improvement in the eGFR slope (*μ*_2_ − *μ*_1_) and its associated uncertainty 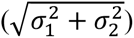 for varying levels of baseline uPCR.
4. Probabilistic Threshold Analysis: To assess clinical significance, we calculated the probability that the eGFR slope improvement would exceed a predefined threshold (e.g., an improvement of >1 ml/min/1.73m² per year).^21,22^ This probability was computed for each patient in the synthetic cohort using the cumulative distribution function (CDF) of the normal distribution for ΔY. The relationship between the percentage of uPCR reduction and the probability of achieving this clinically meaningful outcome was then visualized and analyzed.

## Results

### Data preparation

We used dataset from the study by Iimori et al. (2018) contained 1,138 samples.^17^ To ensure the reliability of the outcome variable, we first selected patients who had five or more eGFR measurements recorded, resulting in a cohort of (n = 655) individuals.^5,19^ From this group, we excluded 119 samples that contained missing values or were considered clinical outliers, defined as having an absolute eGFR slope of 15 ml/min/1.73m²/year or greater. This selection process, detailed in the flowchart in Figure 1A, yielded a final analytical cohort of 536 patients. This cohort was then randomly partitioned into a training set of 428 patients (80%) and a validation set of 108 patients (20%).

**Figure 1.**
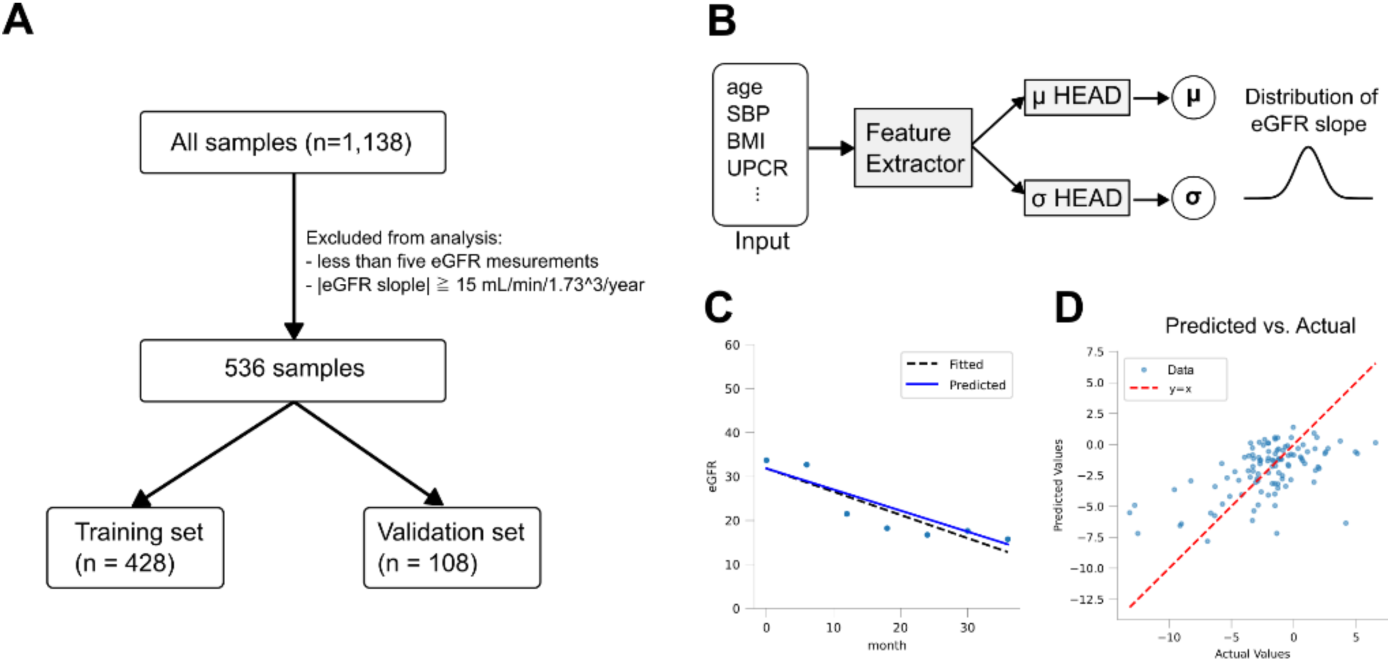
Data preparation and Prediction model. **(A)** Flowchart of the data extraction process used for analysis **(B)** Model architecture for predicting the distribution of eGFR slope values. Predicts the eGFR slope over a 36-month period based on baseline information (month 0). **(C)** Example of eGFR slope prediction for a single sample. Dashed line: fitted line; blue line: predicted line **(D)** Comparison of fitting results and prediction results for validation data. Each point represents an individual sample.

### Creating a model to predict eGFR slope

We developed a probabilistic neural network model to predict the eGFR slope using baseline clinical characteristics (Table 1).^18^ The model architecture, illustrated in Figure 1B, was designed to output a Gaussian probability distribution (defined by a mean, μ, and a standard deviation, σ) for each patient. This approach allows the model to capture not only the most likely rate of eGFR decline but also the uncertainty associated with that prediction. An example of the model’s predictive output for a single patient from the validation set is shown in Figure 1C, illustrating the fitted slope and predicted slope.

**Table 1.** Available Features.

|  | **n** | **mean** | **std** | **min** | **25%** | **50%** | **75%** | **max** |
| --- | --- | --- | --- | --- | --- | --- | --- | --- |
| **gender** | 536 | 1.30 |  | 1 | 1 | 1 | 2 | 2 |
| **age** | 536 | 66.60 | 13.34 | 22 | 60.75 | 69 | 76 | 89 |
| **SBP** | 536 | 138.61 | 21.48 | 69 | 124 | 136 | 150.25 | 234 |
| **BMI** | 536 | 23.96 | 3.79 | 14.24 | 21.28 | 23.60 | 26.38 | 38.37 |
| **etiology of**<br>**CKD** | 536 | 2.32 |  | 1 | 2 | 2 | 3 | 4 |
| **Hb** | 536 | 12.42 | 2.07 | 5.9 | 11.0 | 12.6 | 13.9 | 17.5 |
| **Alb** | 536 | 3.97 | 0.56 | 1.5 | 3.7 | 4.1 | 4.3 | 5.2 |
| **urinary**<br>**occult blood** | 536 | 0.30 |  | 0 | 0 | 0 | 1 | 1 |
| **uPCR** | 536 | 1.59 | 2.70 | 0.00 | 0.10 | 0.53 | 1.79 | 20.18 |
| **hypertension** | 536 | 0.90 |  | 0 | 1 | 1 | 1 | 1 |
| **prevalence**<br>**of CVD** | 536 | 0.24 |  | 0 | 0 | 0 | 0 | 1 |
| **diabetes** | 536 | 0.33 |  | 0 | 0 | 0 | 1 | 1 |
| **use of**<br>**RAASi** | 536 | 0.67 |  | 0 | 0 | 1 | 1 | 1 |
| **use of CCB** | 536 | 0.50 |  | 0 | 0 | 0.5 | 1 | 1 |
| **use of**<br>**diuretics** | 536 | 0.29 |  | 0 | 0 | 0 | 1 | 1 |
| **eGFR** | 536 | 35.79 | 17.34 | 2.47 | 22.22 | 33.45 | 48.43 | 89.98 |
| **eGFR slope** | 536 | -1.75 | 3.19 | -13.30 | -3.33 | -1.63 | -0.04 | 11.43 |

gender: (1: male, 2: female), SBP: Systolic Blood Pressure, etiology of CKD: (1: diabetic, 2: nephrosclerosis, 3: glomerulonephritis, 4: other), CVD: Cardiovascular disease, RAASi: Renin-Angiotensin-Aldosterone system inhibitors, CCB: Calcium Channel Blocker

**Table 2.**
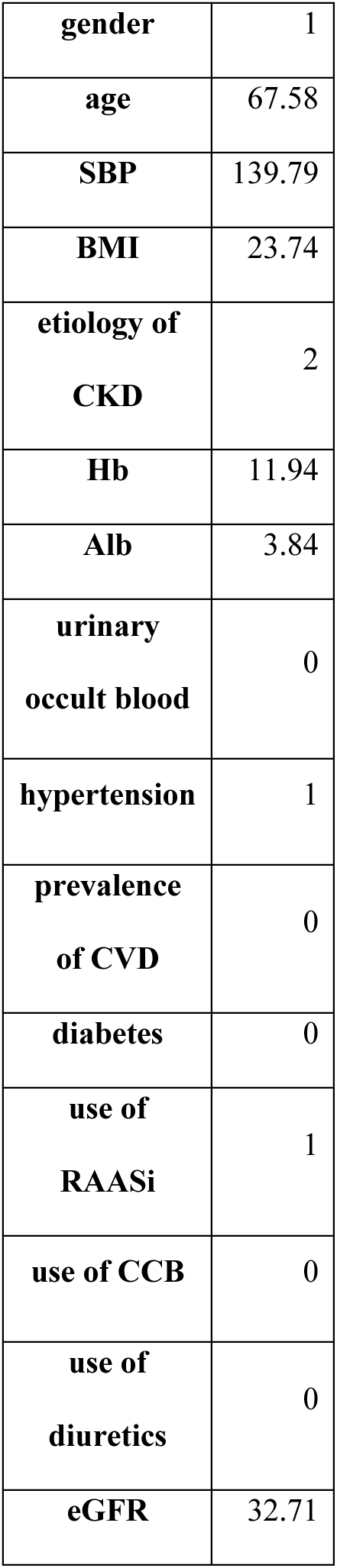
Baseline patients in simulation study.

|  |  |
| --- | --- |
| **gender** | 1 |
| **age** | 67.58 |
| **SBP** | 139.79 |
| **BMI** | 23.74 |
| **etiology of**<br>**CKD** | 2 |
| **Hb** | 11.94 |
| **Alb** | 3.84 |
| **urinary**<br>**occult blood** | 0 |
| **hypertension** | 1 |
| **prevalence**<br>**of CVD** | 0 |
| **diabetes** | 0 |
| **use of**<br>**RAASi** | 1 |
| **use of CCB** | 0 |
| **use of**<br>**diuretics** | 0 |
| **eGFR** | 32.71 |

The model’s performance was evaluated on the hold-out validation set. As shown in the scatter plot in Figure 1D, the predicted mean eGFR slopes demonstrated a strong correlation with the actual calculated slopes. The model achieved a Pearson’s correlation coefficient of 0.56 and a Root Mean Squared Error (RMSE) of 2.81 ml/min/1.73m²/year.

### Analysis of Important Features

To interpret the trained model and identify the key clinical drivers of eGFR slope prediction, we employed the SHAP (SHapley Additive exPlanations) framework. The global importance of each feature, quantified by its mean absolute SHAP value, is presented in Figure 2A. This analysis revealed that the uPCR was the most influential predictor of eGFR slope, followed by Alb and eGFR at month 0.

**Figure 2.**
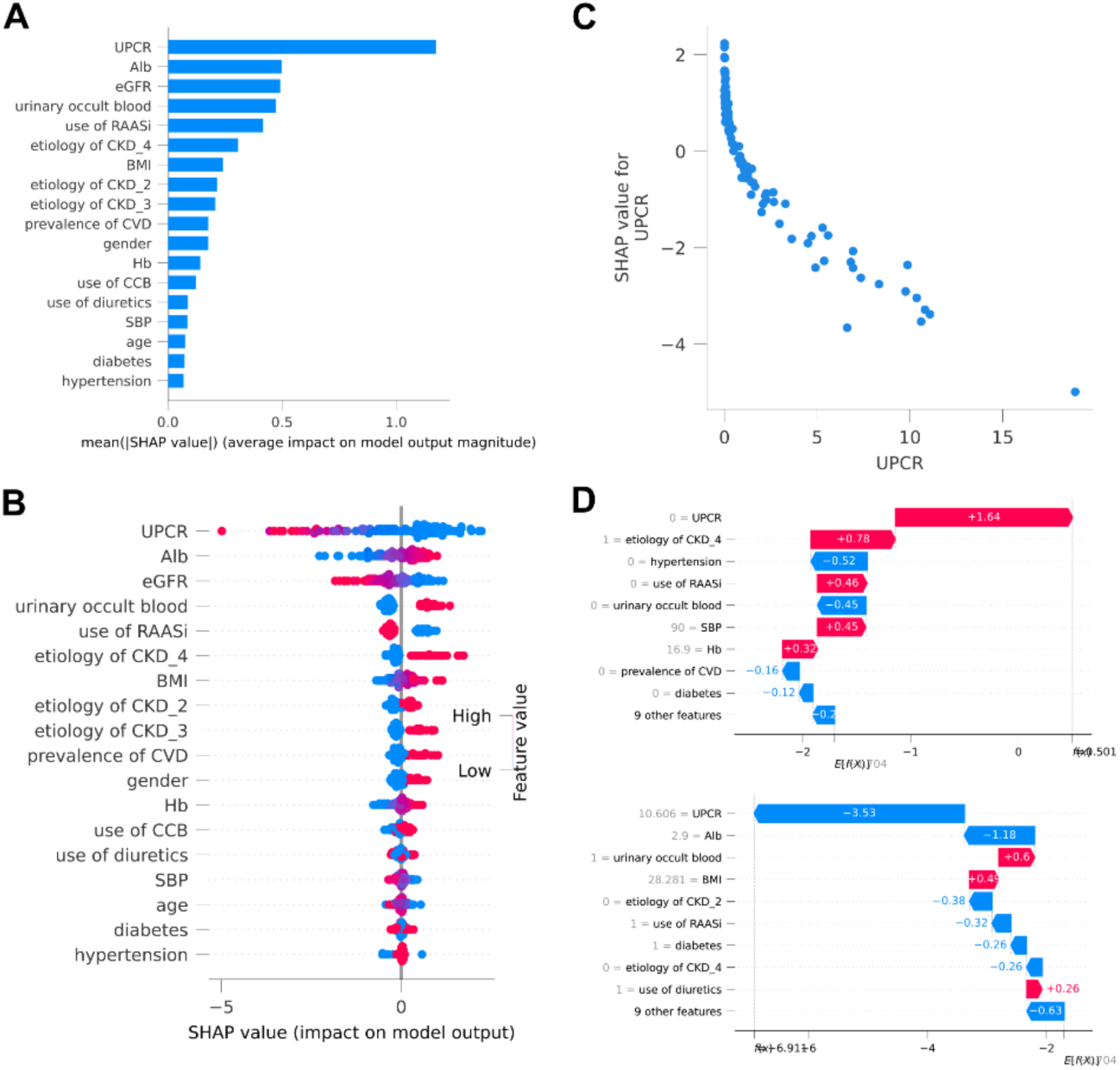
Analysis of important features. **(A)** Feature importance, calculated as the average absolute value of SHAP values. **(B)** SHAP values for each feature, representing their contribution to the model’s predictions. Each point corresponds to a single sample. The color bar indicates the magnitude of each feature’s value. **(C)** Relationship between uPCR values and SHAP values. **(D)** A visualization showing how each feature contributed to the model’s predictions for two specific sample cases.

The SHAP summary plot (Figure 2B) provides a more detailed view of feature contributions, showing both the magnitude and direction of their impact. For uPCR, higher values (indicated by warmer dots) were consistently associated with large negative SHAP values, signifying a strong contribution to predicting a more rapid decline in eGFR (i.e., a more negative slope). The specific relationship between uPCR values and their impact on the model’s output is further detailed in the SHAP dependence plot (Figure 2C), which confirms a clear trend where increasing uPCR levels correspond to more negative SHAP values. To illustrate the model’s decision-making process at an individual level, Figure 2D presents waterfall plots for two representative cases, deconstructing how each feature contributes to the final prediction for those specific patients.

### Simulation study

To quantify the potential clinical benefit of lowering uPCR, we conducted a simulation study using a synthetic patient cohort, as outlined in Figure 3A. We simulated a 62% reduction in uPCR—a clinically relevant intervention target from phase 2 trial of Sibeprenlimab—and used our trained model to predict the eGFR slope before and after this hypothetical change.^20^ An example of this simulation for a single synthetic patient is visualized in Figure 3B, showing a clear positive shift in the predicted eGFR slope distribution after uPCR reduction.

**Figure 3.**
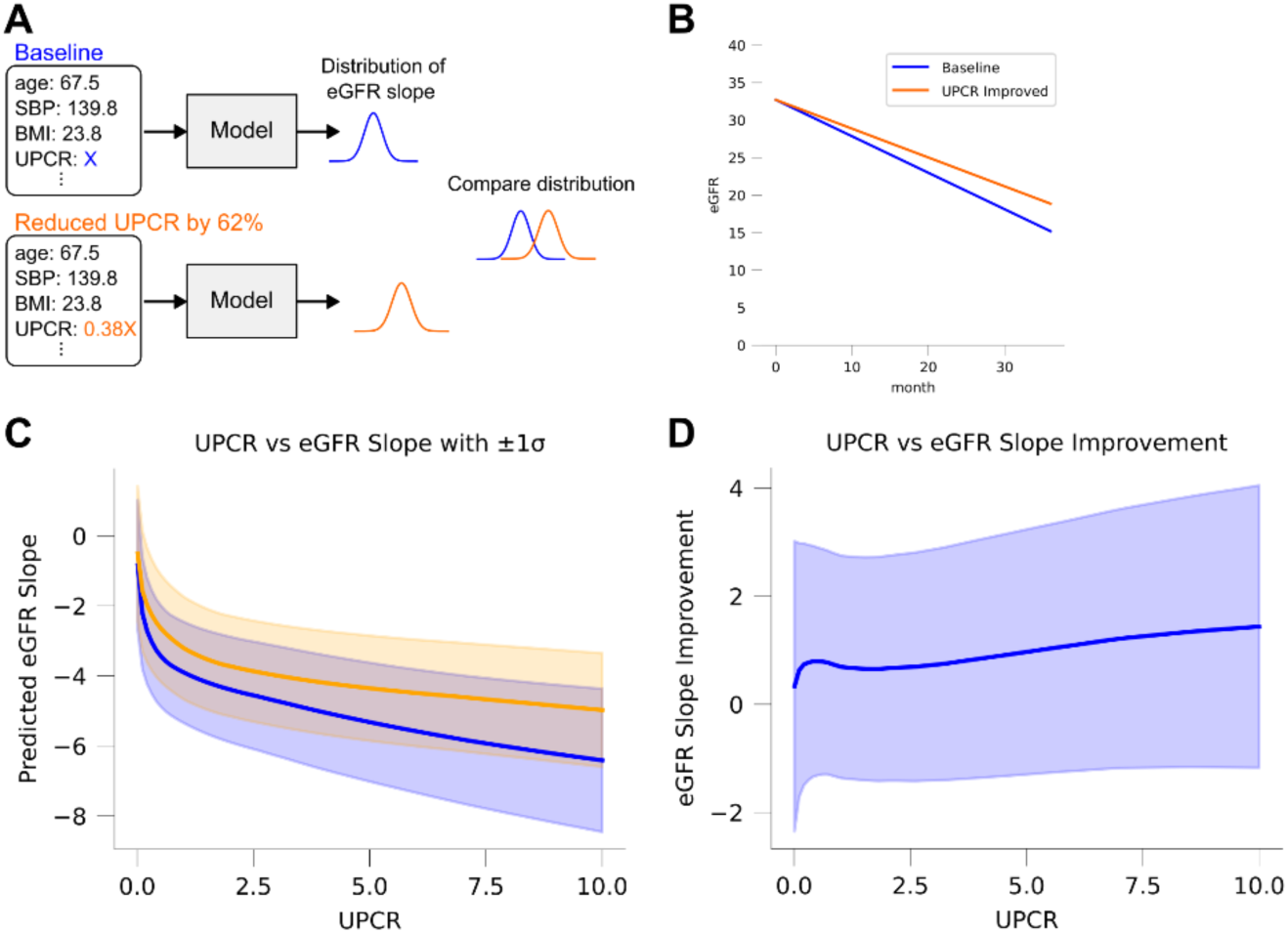
Simulation study for improving uPCR by 62%. **(A)** Overview of the simulation study. We model the distribution of eGFR slope for both the baseline sample and a sample with 62% reduced uPCR, then compare the distributions. **(B)** Example comparison of predicted eGFR slope values before and after reducing uPCR in a specific sample. **(C)** Comparison between baseline uPCR values and corresponding predicted eGFR slope values. Blue: baseline values; orange: 62% reduced uPCR. The shaded area represents the ±1σ confidence interval. **(D)** Relationship between baseline uPCR values and the magnitude of eGFR slope improvement. The shaded area indicates the ±1σ confidence interval.

Across the entire synthetic cohort, the intervention resulted in a consistent and significant improvement in the predicted eGFR slope (Figure 3C). The magnitude of this improvement was found to be dependent on the patient’s baseline uPCR level. As shown in Figure 3D, patients with higher baseline uPCR values experienced a greater predicted improvement in their eGFR slope. For a patient with a baseline uPCR of 7.1, the model predicted an eGFR slope improvement of 1.2 ml/min/1.73m²/year (Figure 3B).

To further explore this relationship, we simulated the effect of varying levels of uPCR reduction on a patient with a high baseline uPCR of 10.0 (Figure 4A). The results, plotted in Figure 4B, demonstrate a clear dose-dependent relationship: greater percentage reductions in uPCR led to progressively larger improvements in the predicted eGFR slope. Specifically, the simulation indicated that to achieve an eGFR slope improvement of 0.5 ml/min/1.73m²/year, a uPCR reduction of 26.2% was necessary, while achieving a more substantial improvement of 1.0 ml/min/1.73m²/year required a 47.4% reduction in uPCR.

**Figure 4.**
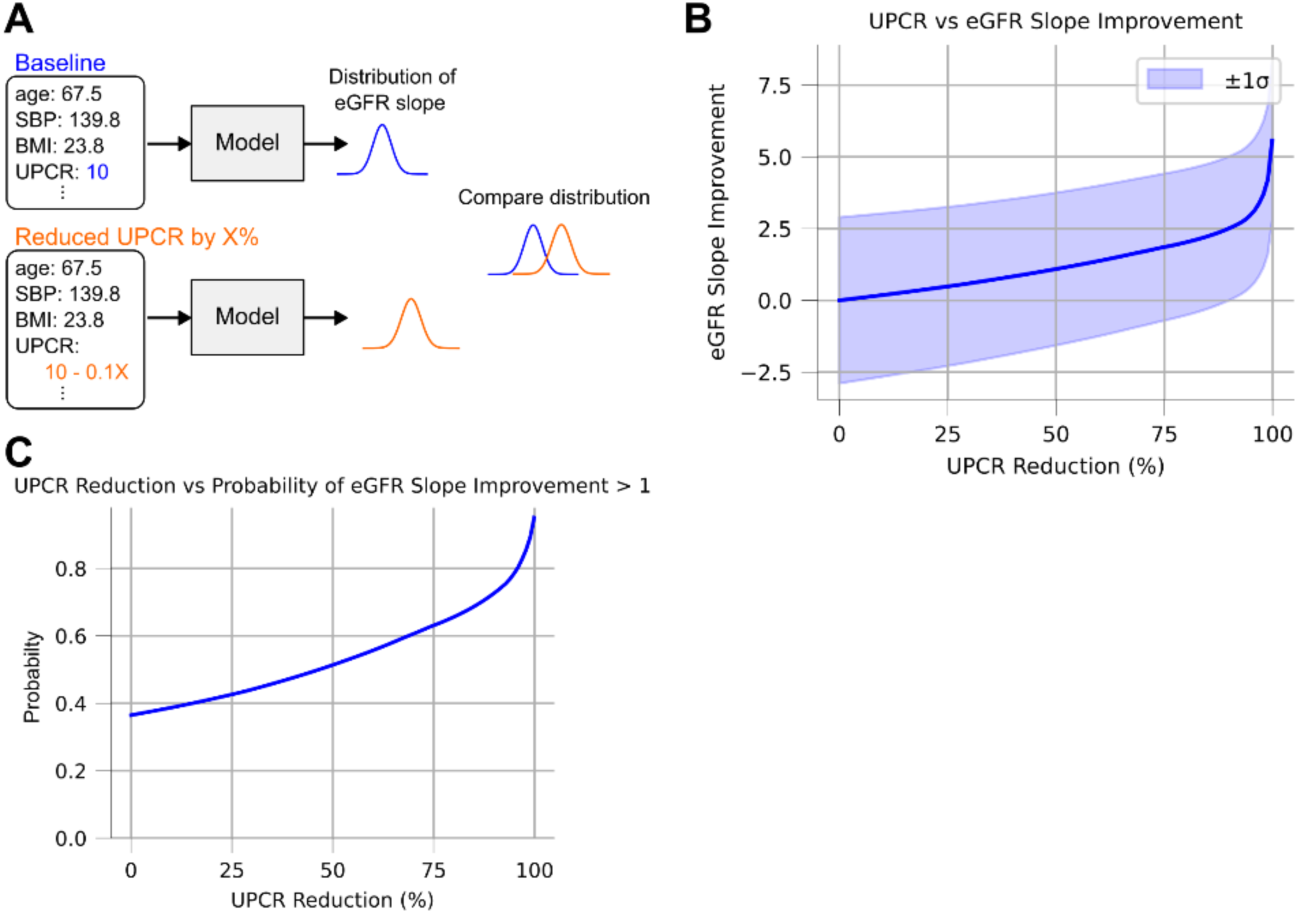
Simulation study for improving uPCR by X%. **(A)** Overview of the simulation study. The baseline sample’s uPCR is set to 10, while the intervention group reduces its uPCR by X% from 10. We predict and compare the distribution of each eGFR slope. **(B)** Relationship between the percentage reduction in uPCR and the improvement in eGFR slope. The shaded area represents the ±1σ interval. **(C)** Relationship between the percentage reduction in uPCR and the probability that the improvement in eGFR slope exceeds 1.0.

Finally, we assessed the clinical significance of this intervention by calculating the probability that the eGFR slope improvement would exceed a clinically meaningful threshold of 1.0 ml/min/1.73m²/year.^19^ Figure 4C shows that this probability increases substantially with the degree of uPCR reduction. For example, a 62% reduction in uPCR from a baseline of 10.0 was associated with a 57% probability of achieving this clinically significant outcome, highlighting the potential impact of effective uPCR management.

## Discussion

In this study, we developed a probabilistic neural network model to predict the long-term eGFR slope and quantify the uncertainty associated with its predictions. We acknowledge the inherent limitations in accurately forecasting future kidney function decline using only baseline clinical information. To address this, our approach of learning a full probability distribution, rather than a single point estimate, allowed for a more nuanced analysis that explicitly considers this uncertainty. Through this lens, our findings reinforce the critical role of uPCR as a key determinant of eGFR slope, suggesting that its reduction contributes to the preservation of kidney function over the medium to long term. Our multi-faceted approach, combining feature importance analysis and a simulation study, provided converging evidence from different perspectives on the significant relationship between uPCR and eGFR slope.

From a technical standpoint, our probabilistic neural network architecture offered several methodological advantages over tree-based machine learning models such as LightGBM.^23^ First, it avoided prediction discontinuities that are often observed in tree-based methods, producing a smoother and more plausible prediction landscape—particularly important in modeling gradual biological processes such as kidney function decline. Second, our feature engineering strategy was critical to the model’s success. Specifically, we included both the raw uPCR values and their logarithmic transformation (log[uPCR]) as model inputs. This allowed the network to learn both linear and non-linear relationships across a wide range of proteinuria levels, thereby enhancing model expressiveness and predictive performance.

However, the use of uPCR reduction as a surrogate endpoint for kidney disease progression is not without its challenges. A decrease in uPCR may, in some cases, reflect a transient pharmacological effect—such as reduced intraglomerular pressure—rather than a true structural improvement or long-term nephroprotective effect.^14,15^ As previously noted, predicting future eGFR slope from a limited set of baseline features remains a formidable challenge.^5,8^ Nevertheless, we believe this study serves as a valuable pilot investigation. By leveraging a publicly available dataset, we have demonstrated the potential utility of uPCR reduction as a valid surrogate endpoint.

This work should be viewed as a proof-of-concept study, laying the groundwork for more extensive analyses. Future research must validate these findings in larger, real-world datasets that encompass patients with diverse backgrounds and treatment histories. Elucidating the precise relationship between uPCR and long-term kidney function in such heterogeneous populations is a critical next step. We believe that this study is a meaningful precursor to such research and will contribute to the design and interpretation of future investigations in the field.

## Data Availability

This study was a secondary analysis of publicly available data from the previous study. (S. Iimori et al. Prognosis of chronic kidney disease with normal-range proteinuria: The CKD-ROUTE study. PLoS One, vol. 13, no. 1, Jan. 2018, doi: 10.1371/journal.pone.0190493.)

## Author Contributions

H.K. supervised the project. A.T. performed the data analysis and prepared the figures. H.I. contributed to figure preparation and wrote the original draft of the manuscript. All authors (A.T., H.I., K.M., Y.Y., H.N., M.O., N.K., H.K.) designed the study. All authors reviewed, provided critical feedback, and edited the manuscript.

